# Gut microbial and human genetic signatures of inflammatory bowel disease increase risk of comorbid mental disorders

**DOI:** 10.1101/2023.12.13.23299882

**Authors:** Junho Lee, Shin Ju Oh, Eunji Ha, Ga Young Shin, Hyo Jong Kim, Kwangwoo Kim, Chang Kyun Lee

**Author notes:** Authors share co-first authorship (Junho Lee, Shin Ju Oh). **Correspondence** Address correspondence to: Kwangwoo Kim, PhD, Kyung Hee University, 26 Kyunghee-daero, Dongdaemun-gu, Seoul 02447, South Korea. or Chang Kyun Lee, MD, PhD, Kyung Hee University College of Medicine, 23 Kyunghee-daero, Dongdaemun-gu, Seoul 02447, South Korea. **Authorship Contributions:** C.K.L. and K.K. designed the research. C.K.L., H.J.K., G.Y.S. and S.J.O. collected the data. J.L., S.J.O., K.K. and C.K.L. analyzed the data, with assistance from E.H. and G.Y.S. J.L., S.J.O., K.K. and C.K.L. wrote the manuscript. All authors contributed to the revision and approval of paper for submission.

## Abstract

The high prevalence of comorbid mental disorders (CMDs), such as anxiety and depression, in patients with inflammatory bowel disease (IBD) is well documented. The reported bidirectional relationship between the two conditions suggests a crucial role of a gut-brain axis in CMD development in patients with IBD. This study aimed to investigate a complex interplay between gut microbiota and host genetic variants relevant to the development of CMDs in IBD. Genome-wide variant data, gut metagenomic data, and/or anxiety/depression estimates were obtained from 507 patients with IBD and 75 healthy controls. A series of integrative analyses were performed, profiling gut microbial diversity, microbial abundance, polygenic risk score, microbial quantitative trait locus (mbQTL), and microbial IBD-risk score. Patients with IBD had significantly lower gut microbial alpha diversity than controls, particularly those with CMD. Beta diversity revealed that a large fraction of IBD-associated taxa contributing to the top principal component were potentially associated with CMD risk. We identified 146 significantly differentially abundant taxa (DATs) between IBD patients and controls, and 48 DATs between CMD-free and CMD-affected IBD patients, with the majority showing consistent changes in abundance between IBD and CMD. Microbial IBD-risk scores, developed to estimate the degree of microbial IBD-specific burden in each individual, supported a significant enrichment of IBD-risk signatures in CMD-affected patients. Additionally, we found an IBD-risk mbQTL for an IBD/CMD-associated DAT, implicating an interplay between IBD-risk variants and gut dysbiosis in the development of both IBD and CMD. Collectively, IBD-associated gut dysbiosis predominantly confers risk of CMD in IBD patients partially through genetic variant-mediated regulation.

## Introduction

Inflammatory bowel disease (IBD), including Crohn’s disease (CD) and ulcerative colitis (UC), causes relapsing and remitting inflammation of the gastrointestinal tract, which has a major impact on the quality of life.^1,2^ Although the etiology of IBD is largely unknown, several risk factors have been identified including genetic variants, immunological cytokines, gut dysbiosis, and environmental triggers.^2-5^

Several epidemiological studies have reported higher frequencies of comorbid mental disorders (CMDs) in patients with IBD. The pooled prevalence of anxiety and depression in patients with inactive IBD has been estimated to be 31.1% and 25.2%, respectively.^6^ These prevalence estimates were much higher than those in the general population (3.8% for anxiety and 3.4% for depression).^7^

Previous reports have provided evidence supporting the association between mental disorders and IBD.^8^ For example, IBD patients with high disease activity exhibited a higher prevalence of anxiety and depression (57.6% and 38.9%, respectively).^6^ Similarly, IBD patients with CMD showed worse prognosis and a high risk of bowel surgery especially in patients with CD.^9,10^ A population-based analysis revealed that depression prior to IBD diagnosis was associated with an increased likelihood of IBD development, especially in individuals with both depression and gastrointestinal symptoms.^11^ In addition, treatment with antidepressants was reported to reduce risk of IBD.^12^ These results indicate that mental disorders do not occur simply due to IBD-related confounding effects on mood (e.g., disease activity, fatigue, flare, frustration, high medical expense).

The accumulated evidence of molecular signatures in both IBD and mental disorders suggested the presence of shared biomarkers in both diseases, suggesting common pathological pathways potentially involved in a bidirectional gut-brain axis.^13-15^ A gut metagenomic study that compared the microbial composition between IBD patients with CMD and those without CMD emphasized abundance changes in a large number of individual taxa according to CMD.^16^ A recent study observed that fecal microbiota transplantation from patients with both IBD and depression causes severe IBD-like colitis and depression-like behavior in mice.^17^ Considering the gut dysbiosis in patients with IBD and the reported gut-brain axis in mental disorders,^18^ it could be worth investigating the correlation of IBD-specific and CMD-specific changes of microbial abundance in a single cohort.

With respect to human genetic factors, a weak but significant genetic correlation was observed between depression and IBD in terms of disease-specific effect sizes of genome-wide variants.^19^ Some of the IBD-risk variants in stress regulator genes were enriched in open chromatin regions interacted with promoters in a brain tissue.^19^ However, the individual genetic variants that explain the pleiotropic mechanism involved in two diseases remain unclear.

In this study, we investigated human genetic variants, gut microbiota, and their interplay that confer risk of CMD in patients with IBD, by profiling the microbial diversity, abundance level, polygenic risk scores (PRS), and microbial quantitative trait loci (mbQTL) in a single-center cohort with a homogenous genetic background.

## Results

### Summary of study design

This study aimed to comprehensively understand the IBD-risk human genetic factors and gut microbial dysbiosis associated with the frequent development of comorbid anxiety and depression in patients with IBD. To this end, we recruited 507 patients with IBD (UC, n = 290; CD, n = 217) and 75 healthy controls to generate genome-wide variant data from peripheral blood, 16S rRNA metagenomic data from fecal samples, and/or Hospital Anxiety and Depression Scale (HADS) data. There were no significant differences in age, body mass index (BMI), cigarette smoking, and alcohol consumption between IBD cases and healthy controls, except sex ratio (**Table 1**). HADS information was obtained from a subset of patients with IBD (n=225). A total of 29 patients with IBD (12.9%) had significant anxiety and/or depression with HADS ≥ 11. Non-psychological characteristics were not significantly different between CMD-free and CMD-affected patients with IBD (**Table 2**).

**Table 1.**
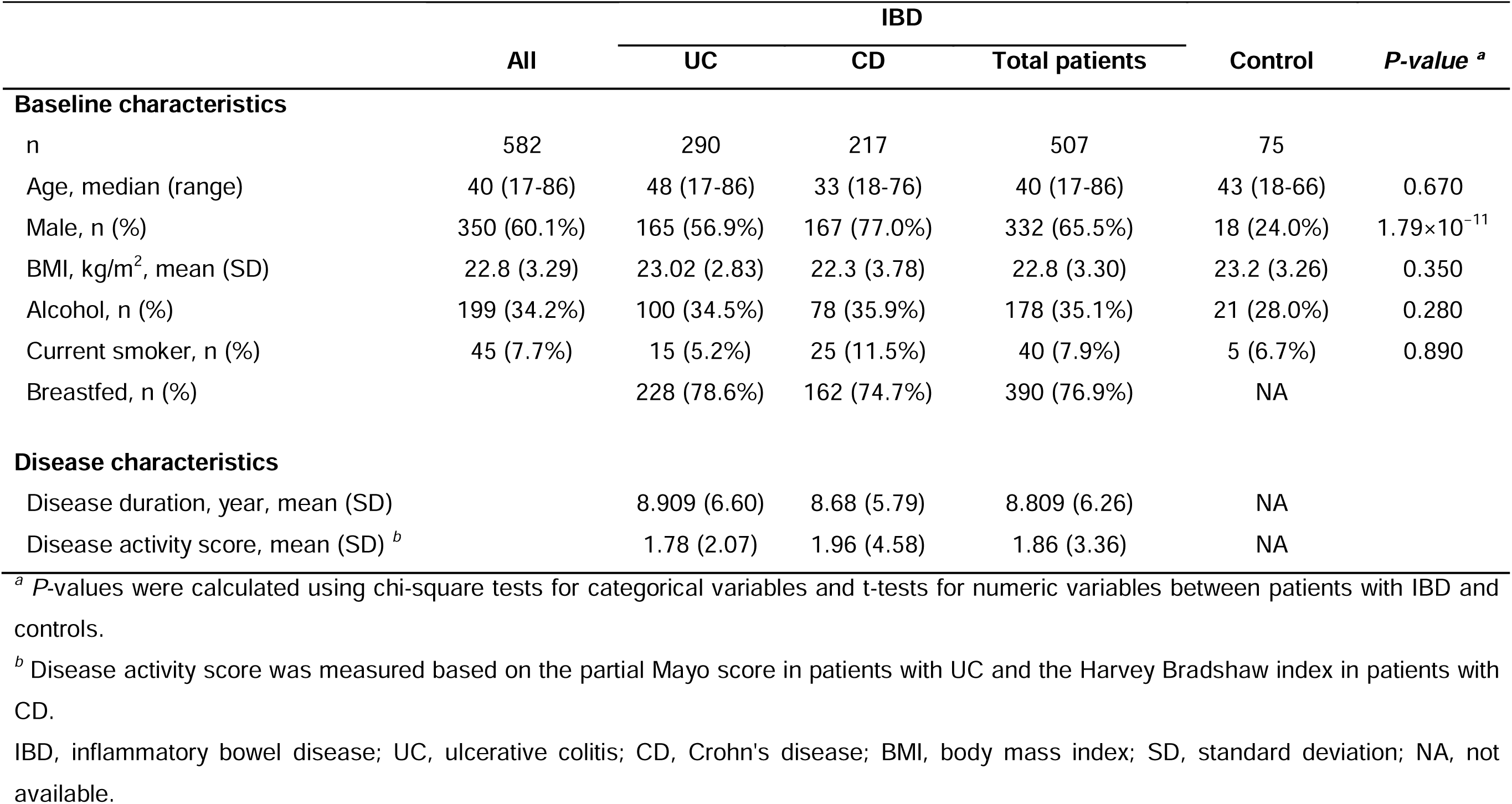
Clinical characteristics of the study participants at a sampling point.

|  | All | IBD |  |  | Control | *P-value*<sup>a</sup> |
| --- | --- | --- | --- | --- | --- | --- |
|  |  | UC | CD | Total patients |  |  |
| Baseline characteristics |  |  |  |  |  |  |
| n | 582 | 290 | 217 | 507 | 75 |  |
| Age, median (range) | 40 (17-86) | 48 (17-86) | 33 (18-76) | 40 (17-86) | 43 (18-66) | 0.670 |
| Male, n (%) | 350 (60.1%) | 165 (56.9%) | 167 (77.0%) | 332 (65.5%) | 18 (24.0%) | 1.79×10<sup>-11</sup> |
| BMI, kg/m<sup>2</sup>, mean (SD) | 22.8 (3.29) | 23.02 (2.83) | 22.3 (3.78) | 22.8 (3.30) | 23.2 (3.26) | 0.350 |
| Alcohol, n (%) | 199 (34.2%) | 100 (34.5%) | 78 (35.9%) | 178 (35.1%) | 21 (28.0%) | 0.280 |
| Current smoker, n (%) | 45 (7.7%) | 15 (5.2%) | 25 (11.5%) | 40 (7.9%) | 5 (6.7%) | 0.890 |
| Breastfed, n (%) |  | 228 (78.6%) | 162 (74.7%) | 390 (76.9%) | NA |  |
| Disease characteristics |  |  |  |  |  |  |
| Disease duration, year, mean (SD) |  | 8.909 (6.60) | 8.68 (5.79) | 8.809 (6.26) | NA |  |
| Disease activity score, mean (SD)<sup>b</sup> |  | 1.78 (2.07) | 1.96 (4.58) | 1.86 (3.36) | NA |  |

<sup>a</sup> *P*-values were calculated using chi-square tests for categorical variables and t-tests for numeric variables between patients with IBD and controls.
<sup>b</sup> Disease activity score was measured based on the partial Mayo score in patients with UC and the Harvey Bradshaw index in patients with CD.
IBD, inflammatory bowel disease; UC, ulcerative colitis; CD, Crohn's disease; BMI, body mass index; SD, standard deviation; NA, not available.

**Table 2.** Clinical characteristics of IBD patients with HADS information at a sampling point.

|  | CMD-free IBD | CMD-affected IBD <sup>a</sup> |  |  | *P-value* <sup>b</sup> |
| --- | --- | --- | --- | --- | --- |
|  | All (n=196) | All (n=29) | Anxiety (n=15) | Depression (n=22) |  |
| Age, median (range) | 37 (18-77) | 35 (21-59) | 30 (22-59) | 38.5 (21-59) | 0.397 |
| Male, n (%) | 135 (68.9%) | 22 (75.9%) | 10 (66.7%) | 17 (77.3%) | 0.584 |
| BMI, kg/m<sup>2</sup>, mean (SD) | 22.8 (3.32) | 22.4 (3.86) | 22.5 (4.48) | 21.9 (3.0) | 0.608 |
| Alcohol, n (%) | 82 (41.8%) | 10 (34.5%) | 5 (33.3%) | 6 (27.3%) | 0.583 |
| Current smoker, n (%) | 15 (7.65%) | 2 (6.9%) | 1 (6.7%) | 1 (4.5%) | 1.000 |
| Breastfed, n (%) | 154 (78.6%) | 20 (69.0%) | 9 (60.0%) | 15 (68.2%) | 0.419 |
| Subtype, n (%) |  |  |  |  |  |
| UC | 82 (41.8%) | 13 (44.8%) | 7 (46.7%) | 9 (40.9%) |  |
| CD | 114 (58.2%) | 16 (55.2%) | 8 (53.3%) | 13 (59.1%) |  |
| Disease activity score, mean (SD) <sup>c</sup> |  |  |  |  |  |
| PMS (for UC) | 1.84 (2.15) | 2.62 (3.38) | 3.71 (4.35) | 3.22 (3.93) | 0.437 |
| HBI (for CD) | 1.32 (3.43) | 2.19 (2.04) | 2.50 (2.07) | 1.77 (1.74) | 0.159 |
| Disease duration, year, mean (SD) | 9.13 (5.79) | 7.39 (3.98) | 8.03 (3.96) | 7.20 (4.17) | 0.0454 |

<sup>a</sup> CMD in patients with IBD was defined as having significant anxiety or depression with a HADS-A/D score of 11 or higher.
<sup>b</sup> *P*-values were calculated using chi-square tests for categorical variables and t-tests for numeric variables between patients with CMD-free and CMD-affected IBD.
IBD, inflammatory bowel disease; CMD, Comorbid mental disorder; BMI, body mass index; SD, standard deviation; UC, ulcerative colitis; CD, Crohn's disease; PMS, partial Mayo score; HBI, Harvey-Bradshaw index.

To dissect the potential causal relationships between IBD-risk factors and mental disorders (depicted in **Figure 1**), we first tested whether IBD-risk microbial signatures, including microbial diversity and taxon-level abundance, were more predominant in IBD patients with CMD than in those without CMD. Second, we tested whether the effects of genome-wide variants on IBD were correlated with those on mental disorders in the general population and whether PRSs for IBD or mental disorders were higher in IBD patients with CMD than in those without CMD. Finally, we tested whether IBD-risk genetic variants mediate changes in the abundance of microbiota that confer risk of CMD.

**Figure 1.**
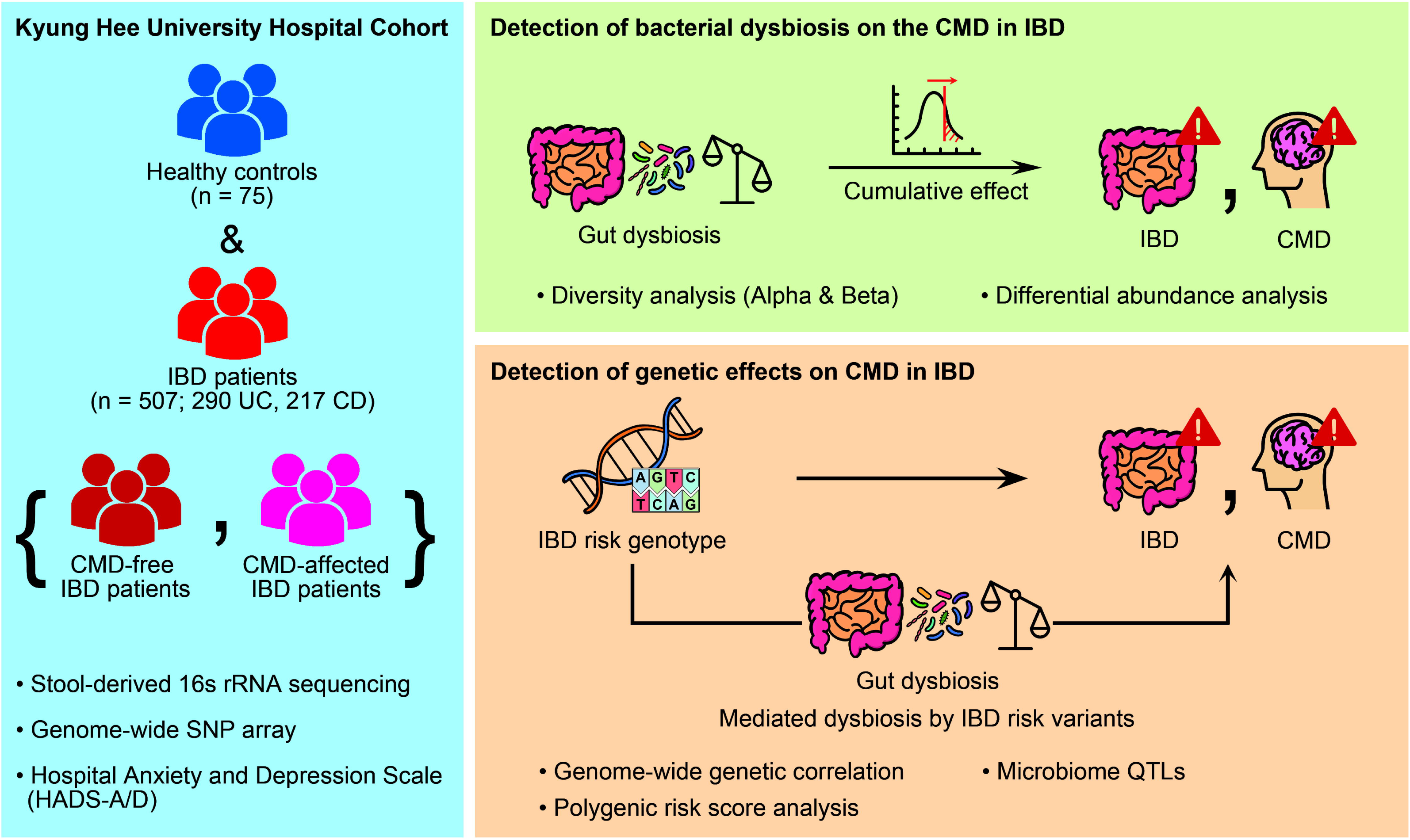
Summary of the study strategy. The figure shows a simplified representation of the research scope and strategies to investigate the contributions of IBD-risk factors to the development of CMD using a large Korean case-control cohort with gut metagenomic and human genetic data and clinical outcomes.

### Enhanced IBD-specific gut dysbiosis in IBD patients with CMD

We estimated gut microbial diversity and genus-level abundance in Korean patients with IBD and healthy controls. Shannon alpha diversity index was significantly lower in patients with IBD (mean alpha diversity 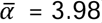) than healthy controls 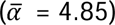 at the genus level (*P* = 3.93×10^−15^; **Figure 2a**), showing similar differences in alpha diversity between each IBD subtype and the control group (*P* = 2.90×10^−11^ for UC and *P* = 6.26×10^−19^ for CD, respectively; **Supplementary Figure S1**). The stratification of patients with IBD according to CMD status revealed that IBD patients with CMD had even less alpha diversity 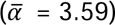 in the gut microbiota compared to controls (*P* = 6.69×10^−09^) or CMD-free IBD patients (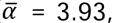 *P* = 0.047; **Figure 2a**). Alpha diversity in the anxiety group 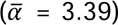 was relatively lower than that in the depression group 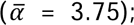 however, the difference was not statistically significant (*P* = 0.189).

**Figure 2.**
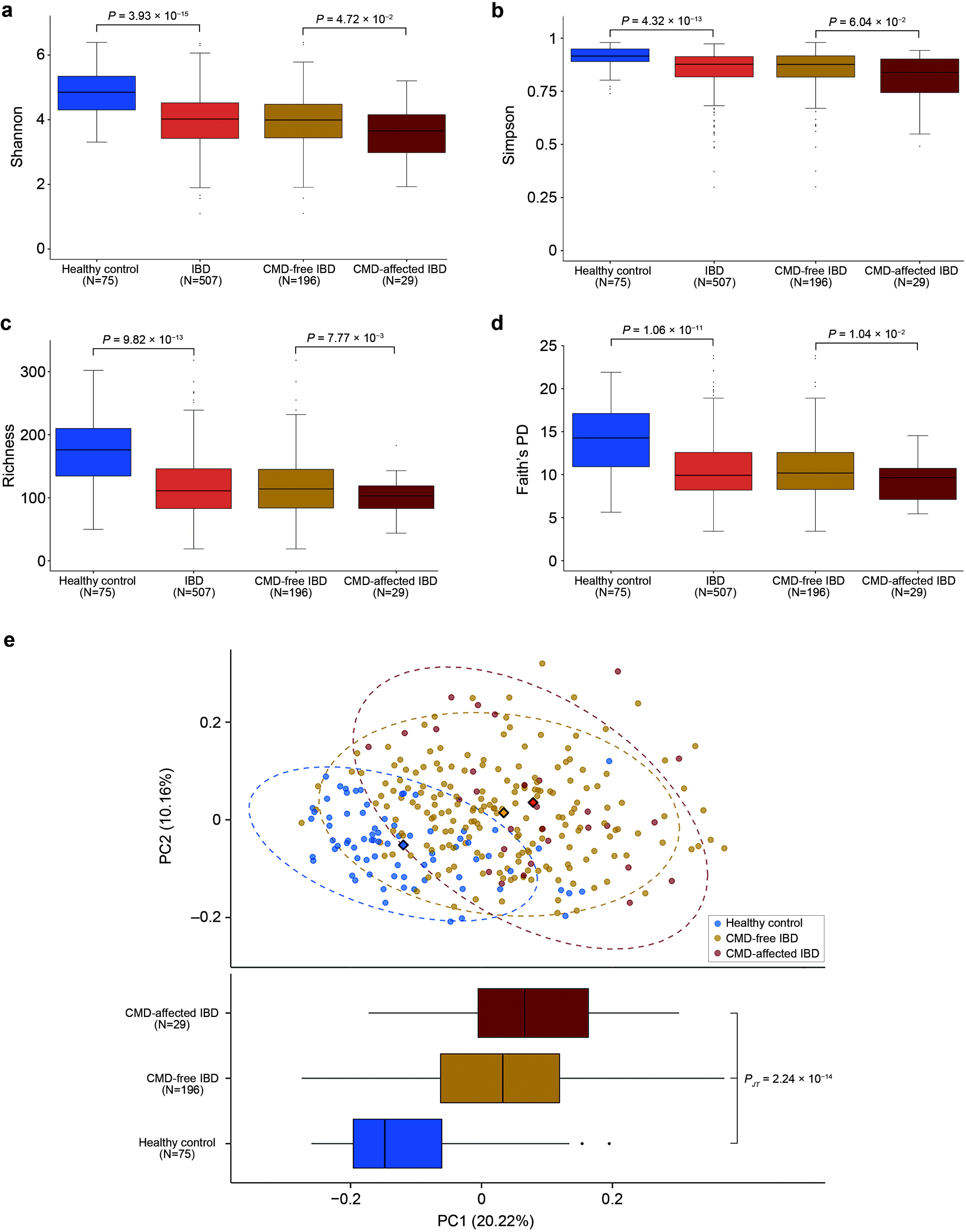
Microbial alpha and beta diversity indices. **a-d,** Alpha diversity was assessed based on the Shannon index (**a**), Simpson index (**b**), richness (**c**), and Faith’s PD index (**d**) at the genus level. P-values for statistical differences in alpha diversity were calculated between healthy controls and patients with IBD and between CMD-free and CMD-affected IBD patients. **e,** Beta diversity was measured based on the Bray-Curtis dissimilarity of individuals’ genus abundance and visualized in a PCoA plot according to the top two principal components (PCs). The centroid of each cluster is marked by a diamond. The top PC that explained the largest proportion (20.22%) of variance in microbial composition in the study subjects is shown in a boxplot according to the affected status. The P-value from a Jonckheere-Terpstra test (*P*_JT_) was calculated to test for a significant trend toward higher PC1 values in the order of healthy controls, CMD-free IBD patients, and CMD-affected IBD patients.

Consistently, other diversity indices were significantly lower in patients with IBD than healthy controls (*P* = 4.32×10^−13^ for Simpson index, *P* = 9.82×10^−13^ for richness, *P* = 1.06×10^−11^ for Faith’ phylogenetic diversity (PD) index), especially in CMD-affected IBD patients than CMD-free IBD patients (*P* = 0.0604 for Simpson index, *P* = 7.77×10^−3^ for richness, *P* = 0.0104 for Faith’s PD index; **Figure 2**). In the same analysis for each subtype (UC and CD) of IBD, we observed a similar trend of decreased alpha diversity in patients with CMD but this did not achieve statistical significance, probably due to the decreased sample sizes in a stratification analysis (**Supplementary Figure S1**).

We then evaluated beta diversity using the individual-to-individual Bray-Curtis dissimilarity from the gut microbiome composition at the genus level. There was a significant dissimilarity of the gut microbiome composition between patients with IBD and healthy controls (*P* ≤ 0.001; **Figure 2e**; see also **Supplementary Figure S2**). In a principal coordinate analysis (PCoA) for the Bray-Curtis dissimilarity, the top principal component (PC1) explained 20.22% of the variance in the dissimilarity of gut microbiome composition among individuals and were significantly higher in patients with IBD than healthy controls (*P* = 8.07×10^−20^; **Figure 2e**). In addition, there was a significant trend toward higher PC1 values in the order of healthy controls, CMD-free IBD patients, and CMD-affected IBD patients (*P* = 2.24×10^−14^ in a Jonckheere-Terpstra test among the three groups, *P* = 0.0345 between CMD-free and CMD-affected IBD patient groups). Thus, it is tempting to suggest that a part of the microbiota that largely contributes to the top PCs may explain the risk of both CMD and IBD.

Given the alpha and beta diversity analysis results, we hypothesized that a part of the IBD-associated gut microbiota is further involved in the development of CMD, which may explain the high prevalence of depression and anxiety in patients with IBD. In this context, we aimed to investigate the taxa whose abundance changes were associated with the risk of both IBD and CMD with the same direction of abundance changes in these two diseases.

We identified 146 differentially abundant taxa (DATs), including seven phyla, 11 classes, 19 orders, 34 families, and 75 genera, between patients with IBD and healthy controls at a false discovery rate threshold of 5% (**Supplementary Table S2**). Of the IBD-associated DATs with complete taxonomic classification information, a total of 18 identified taxa (24.3%) have already been reported to exhibit differential abundance in previous IBD gut metagenomic studies at various taxonomic levels,*^20,21^* showing the consistent directions of abundance changes in patients with IBD; they included the phyla Firmicutes, Tenericutes; classes Clostridia, Gammaproteobacteria, and Mollicutes; the order Clostridiales; the families *Enterobacteriaceae*, *Enterococcaceae*, and *Mogibacteriaceae*; the genera *Barnesiella*, *Bilophila*, *Butyricimonas*, *Campylobacter*, *Coprococcus*, *Desulfovibrio*, *Eggerthella*, *Paraprevotella*, and *Pediococcus*. In addition, a large number of IBD-associated DATs were newly identified by taking advantage of using a large-scale IBD cohort with an underrepresented ethnicity (**Supplementary Table S2**). Considering the ancestral heterogeneity and the significant number of replicated associations of known IBD-associated taxa, novel IBD-associated DATs are likely to be useful biomarkers and therapeutic targets, especially in East Asian patients with IBD.

In parallel, we identified the significantly differential abundance of 48 taxa between CMD-free and CMD-affected patients with IBD (**Supplementary Table S2**). Of the identified DATs with complete taxonomic classification information, 17 taxa (35.4%) have been associated with anxiety or depression in previous studies.^16,22-25^ The CMD-associated DATs were significantly enriched in IBD-associated DATs (*P* = 1.82×10^−3^; odds ratio=2.66 with 95% confidence interval from 1.39 to 5.14). Specifically, of the 146 IBD-associated taxa, 26 showed significant differences in abundance between the CMD-affected and CMD-free IBD patient groups. Of these 26 DATs, 23 taxa exhibited the same direction of abundance changes between patients with IBD and healthy controls and between CMD-affected and CMD-free patients with IBD (**Figure 3**; see also **Supplementary Table S2**). This result indicated that patients with CMD-affected IBD demonstrated a significantly greater deviation in the abundance levels of these taxa from those observed in healthy controls, in comparison to the degree of deviation exhibited by patients with CMD-free IBD from controls. We also noted that 33 IBD-associated genera, even without statistical evidence of CMD associations, showed consistent directions of abundance changes in IBD and CMD (**Figure 4**).

**Figure 3.**
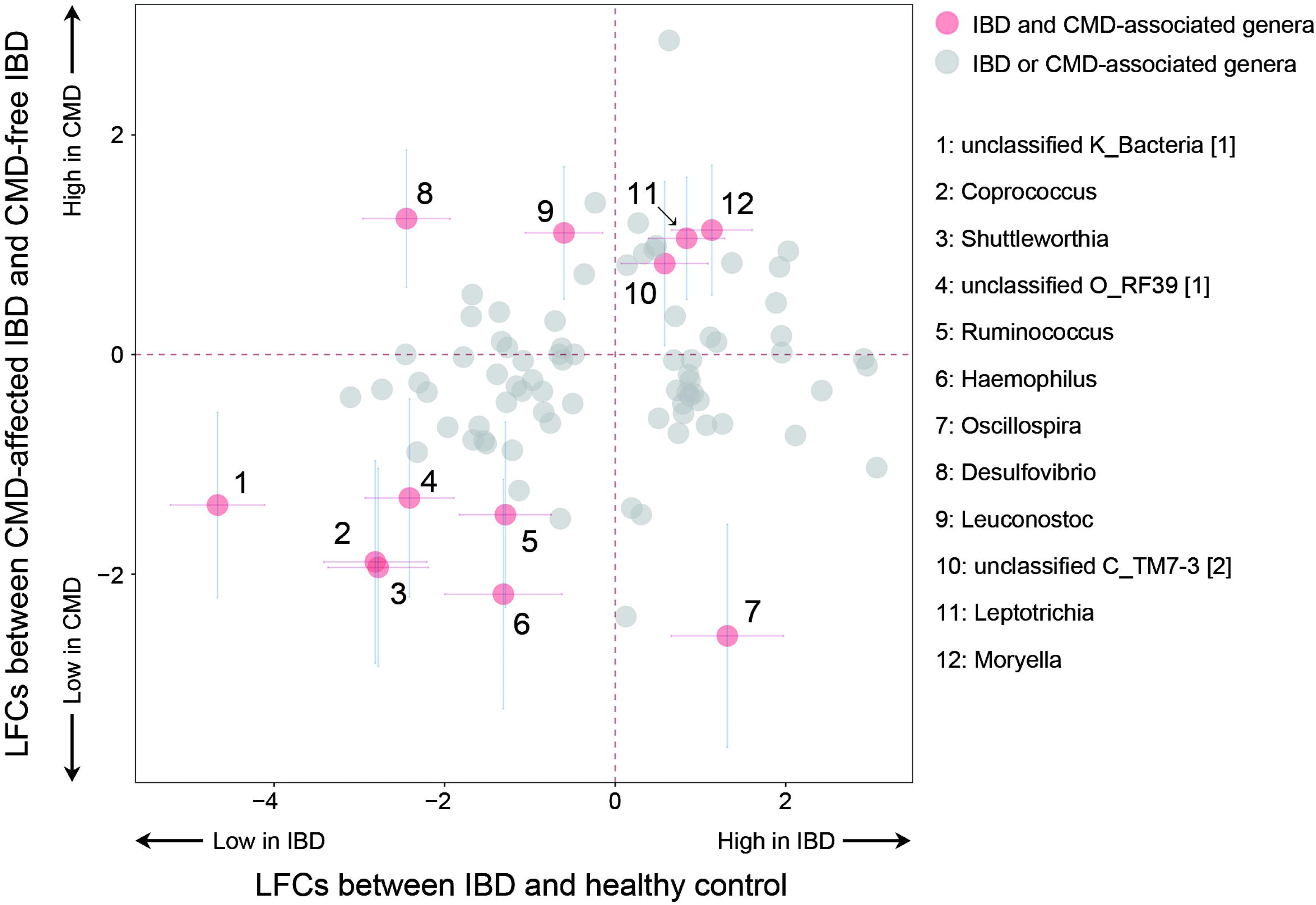
Comparison of the microbial abundance changes specific to the status of IBD and CMD. Log_2_-fold changes (LFCs) in microbial abundance were estimated at the genus level in a differential abundance analysis between healthy controls and patients with IBD and between CMD-free and CMD-affected IBD patients. The scatter plot represents phenotype-specific LFCs for genera associated with either IBD or CMD (*P*_FDR_<0.05). Data points highlighted in red with error bars of 95% confidence intervals indicate the genera whose abundance changes were significant in both phenotypes. Their names were provided with the initial of the lowest taxonomic rank that was classified.

**Figure 4.**
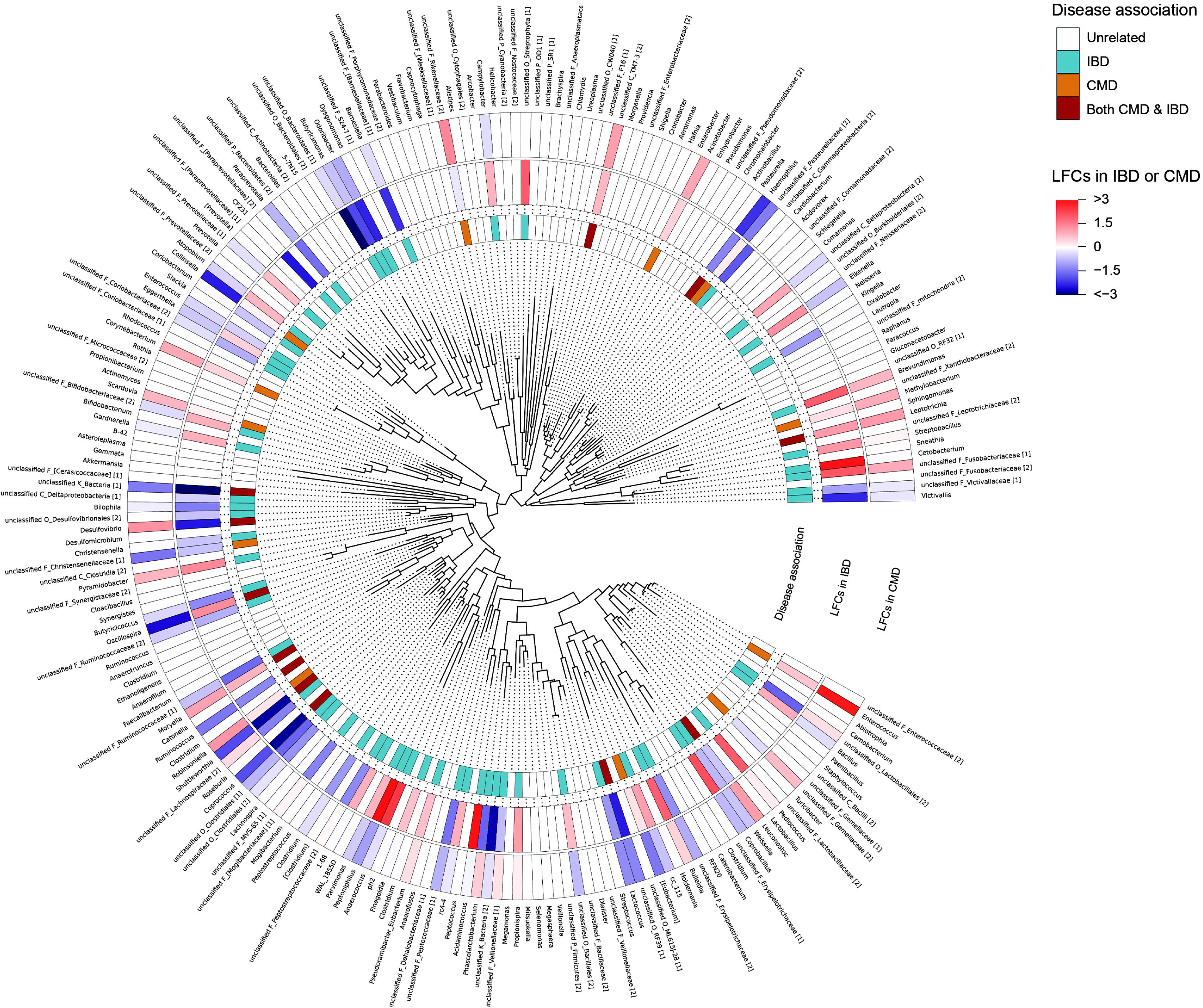
Overview of the gut microbiome and disease-specific abundance changes. A phylogenetic tree of the gut genera detected in this study was constructed based on the FastTree distance between the ASV sequences. The single most abundant ASV of each genus was used in the phylogenetic analysis. The inner heatmap provides information on the phenotypes associated with each genus (*P*_FDR_<0.05). The middle and outer heatmaps log_2_- fold changes (LFCs) in abundance changes in IBD (compared to the control) and CMD-affected IBD (compared to CMD-free IBD), respectively. The name of unclassified genera starts with “unclassified” followed by the initial of the lowest taxonomic levels specified (e.g., unclassified C_TM7-3). Unclassified taxa with different ASVs but sharing the same name are distinguished by numbers in square brackets at the end of their respective names.

To assess whether the CMD-specific microbial change enriches the microbial risk factors for IBD, we estimated the IBD-specific microbial burden as a single statistic (microbial risk score; MRS), which is a linear summation of the normalized abundance levels of CMD-associated genera in an individual weighted by the pre-estimated IBD-specific effect sizes (see the Methods section). We found that an increased MRS for IBD is associated with an increased risk of CMD in patients with IBD (*P* = 1.44×10^−3^). When the study subjects were dichotomized into two groups at the median MRS, the high-MRS group had a large odds ratio of 12.6 (95% confidence interval = 2.9 to 55.6) for CMD development in a multivariate logistic regression (**Table 3**).

**Table 3.** Increased risk of CMDs in individuals with high IBD-specific MRS.

|  | IBD-specific MRS |  | Association between the high MRS group and CMD development |  |  |  |
| --- | --- | --- | --- | --- | --- | --- |
|  |  |  | vs. Control |  | vs. CMD-free IBD |  |
|  | High group, n (%) | Low group, n (%) | OR (95% CI) | P-value | OR (95% CI) | P-value |
| Control | 20 (26.7%) | 55 (73.3%) | 1 (reference) | - | - | - |
| CMD-free IBD | 103 (52.6%) | 93 (47.4%) | 3.3 (1.7 to 6.4) | $3.16 \times 10^{-4}$ | 1 (reference) | - |
| CMD-affected IBD | 27 (93.1%) | 2 (6.9%) | 73.7 (10.0 to 543.5) | $2.44 \times 10^{-5}$ | 12.6 (2.9 to 55.6) | $7.85 \times 10^{-4}$ |

Odds ratios (ORs) and 95% confidence intervals (CIs) were calculated using logistic regression, adjusting for potential confounding factors included in the differential abundance analysis. MRS, microbial risk score; IBD, inflammatory bowel disease; CMD, comorbid mental disorders.

### No evidence of IBD-risk genetic variants on the susceptibility to CMD in patients with IBD

Genome-wide association studies have identified many risk loci for IBD^26,27^ and mental disorders^28,29^. A positive correlation between the disease-risk effects of genome-wide genetic variants on IBD and mental disorders has been previously reported using LD score regression (LDSC).^19^ Thus, it is likely that a high genetic burden from the reported IBD-risk variants increases the susceptibility to CMD in patients with IBD.

To test this hypothesis, we calculated individual disease-specific PRSs based on the reported disease effect sizes of genetically independent risk variants (n=136 for IBD, 87 for UC, 119 for CD, 70 for anxiety, or 49 for depression) from previous genome-wide association studies (GWASs) in multiple populations^26-29^, as described in the Methods section. We note that the previously reported significance of the LDSC-based genetic correlation between IBD and mental disorders was replicated in the public GWAS results used in our PRS analysis (**Supplementary Table S3**).

In a combined analysis with additional out-of-study controls, the patients with IBD showed significantly higher disease-specific PRSs for IBD, CD, UC, and depression than those of the controls (8.17×10^−13^ ≤ *P* ≤ 0.0182; **Supplementary Figure S3**) except for anxiety. However, there was no evidence of the involvement of the genetic burden of IBD-risk variants in the development of CMD (*P* = 0.641). Moreover, patients with CMD did not show significantly higher PRSs for anxiety and depression than those without CMD (*P* ≥ 0.321). This result indicates a relatively weak genetic effect of mental disorder-risk variants on the development of CMD in patients with IBD or suggests that the development of CMD in IBD may have a different etiology from the general mental disorders that develop in the general population.

### Regulatory effects of IBD-risk variants on CMD-risk microbial abundance changes

We searched for mbQTLs for each CMD-risk taxon through a genus-wide case-only GWAS using both metagenomic and genetic data from patients with IBD, followed by a meta-analysis with a previous mbQTL from a larger-scale study^30^ involving multiple cohorts. We evaluated the genetic overlap of mbQTLs for CMD-risk taxa with known risk variants for IBD and mental disorders.

Among the 25 taxa associated with both IBD and CMD, we identified that the T allele of rs35866622 in the FUT2-FUT1 locus was associated with the decreased abundance of the genus *Ruminococcus* (*torques* group; *P* = 2.43×10^−8^, β=−0.061, 95% confidence interval= −0.082 to −0.040; **Supplementary Figure S4**). The genus *Ruminococcus* is a well-known IBD-associated taxon^31^ and our study showed a significant decrease in its abundance in both patients with IBD (compared to controls) and patients with CMD-affected IBD (compared to patients with CMD-free IBD). In addition, the identified mbQTL rs35866622 for the genus *Ruminococcus* has been reported as a lead disease-risk variant of IBD and CD, and its T allele conferred risk of the diseases.^26,27^ Therefore, it is plausible to suspect that the IBD-risk allele in the FUT2-FUT1 locus decreased the abundance of the genus *Ruminococcus*, thus potentially increasing the genetic liability to both IBD and CMD.

The locus around rs35866622 contains a number of proxy variants of the lead variant in a high LD, including a nonsense variant rs601338 in FUT2 (*r*^2^ = 1.00 and 0.80 in 1KGP East Asians and 1KGP Europeans, respectively). FUT2 encodes α-(1, 2)-fucosyltransferase enzymes responsible for the secretion of H antigens in body fluids, including the gut mucosa. The absence of H antigens in the gut mucosal fluid, caused by two copies of nonfunctional FUT2 alleles with the nonsense allele of rs601338, has been reported to have a large impact on the abundance levels of several taxa,^30,32^ some of which have been previously associated with IBD-associated dysbiosis.^30,32,33^

## Discussion

This is the first study to investigate both host genetic and gut microbial factors in CMD-affected IBD patients in a single cohort. Our integrative analysis of the human genome and gut metagenome revealed that IBD-risk dysbiosis was significantly enhanced in patients with CMD, whose gut microbiota was characterized by less diversity and more dysregulated abundance levels of IBD-associated microbial taxa than those without CMD. An MRS analysis showed that an enrichment of microbial IBD-risk factors dramatically increases susceptibility to CMD. Moreover, an IBD-risk mbQTL allele decreases the abundance of the genus *Ruminococcus* that is protective against both IBD and CMD. Our results strongly suggest that the gut-brain axis plays a crucial role in the development of CMD in patients with IBD, with IBD-specific gut dysbiosis primarily contributing to CMD risk and a host-gut microbiota interaction partially mediating this relationship.

We identified nine genera associated with the risk of both IBD and CMD, with the same direction of abundance changes in both conditions (**Figure 3**). This indicates that extremely severe alteration in the abundance of IBD-associated taxa in patients with CMD-affected IBD. For instance, the genus *Coprococcus* decreased 7.0-fold in patients with IBD compared to healthy controls and 3.7-fold in patients with CMD compared to those without CMD. Indeed, the same genus was 15.4-fold lower in CMD-affected IBD patients than in healthy controls (**Supplementary Table S2**).

The genus *Coprococcus* is a butyrate-producing microbiome involved in the hypothalamic-pituitary-adrenal axis and is known to be depleted in individuals with depression and stress.^18,34^ The reduced production of short-chain fatty acids, including butyrate, facilitates inflammation or barrier dysfunction in the gastrointestinal tract.^35,36^ In addition, a recent study suggested a possible role of butyrate-unrelated mechanisms in *Coprococcus* in association with quality of life.^18^

Another important observation was that the IBD-risk variant rs35866622 was an mbQTL for CMD-associated bacteria (**Supplementary Figure S4**). We examined the genetic effects on the compositional changes in the microbiome to understand the mediating effect between IBD-risk variants and CMDs through alterations in the microbiome. The IBD-risk mbQTL is suggested to regulate the IBD/CMD-associated genus *Ruminococcus* by regulating the secretion of H antigens in the gut mucosa.^32,37,38^ This finding suggests that the genetic burden of IBD partially contributes to the frequent occurrence of mental disorders in patients with IBD, suggesting that FUT2 proteins and H antigens might be potential therapeutic targets for IBD and CMD. *Ruminococcus* spp., butyrate-producing bacteria that are abundant in the gut microbial community,^39,40^ were also decreased in patients with IBD in our study, especially in patients with CMD. A similar decrease in *Ruminococcus torques* was previously reported in patients with CD.^41^ A discussion of the other IBD/CMD-associated genera is provided in **Supplementary Note 1**.

Notably, CMD-associated taxa without significant IBD associations need to be further investigated to understand whether they are IBD-independent mental disorder-risk factors or secondary outcomes from mental disorder-mediated effects on microbiome composition. For example, we found a CMD association of the order Coriobacteriales, the families *Coriobacteriaceae* and *Christensenellaceae*, and the genera *Collinsella*, *Enterococcus*, and *Eubacterium*, which were previously reported to be associated with anxiety or depression in the same direction as abundance changes in affected individuals (**Supplementary Table S2**).

Although a significant but weak genetic correlation between mental disorder in the general population and IBD was observed in an analysis using large-scale public GWAS results (**Supplementary Table S3**), the PRS-level genetic burden for IBD or mental disorders was not significantly different between CMD-affected and CMD-free IBD patients in our study cohort (**Supplementary Figure S3**). Given the small sample size for the genetic association analysis, our results indicate that pleiotropic variants for the risk of IBD and mental disorders have little or much weaker effects on the development of CMD in patients with IBD than the effects of gut dysbiosis on CMD.

A case-only study in Switzerland was previously conducted to identify DATs in the gut correlated with HADS-A or HADS-D scores in patients with IBD in remission.^16^ Among the reported CMD-associated genera with complete classification information (n = 14), the CMD association of three genera was replicated in our Korean IBD cohort, showing the largest abundance changes between CMD-free and CMD-affected patients with IBD (**Supplementary Table S2**). Other inconsistent results may indicate ancestry-specific heterogeneity in microbial effects on CMDs between European and East Asian populations, possibly due to the population-specific gut microbial community and the population-specific abundance levels of CMD-associated taxa that could be associated with genetic background, diet, and lifestyle, as well as the degree of statistical power in a differential microbial abundance analysis. In addition, the Swiss sample size (n = 204) was moderate and was further stratified in the differential abundance analysis according to the type of IBD or CMD. Additionally, the statistical models used in the two studies differed (linear regression vs. generalized linear regression using a negative binomial distribution).

This study provided unique opportunities to examine the gut metagenome and human genome-wide SNP data simultaneously in a single center cohort, which allowed us to investigate gut dysbiosis and human genetic data separately, as well as their inter-omics connections. In addition, conducting both case-control and case-only metagenomic analyses within the same cohort could offer more reliable identification of shared risk factors between IBD and CMD, as compared to conducting separate analyses using two independent cohorts with different conditions such as genetic background, diet, and disease activities.

Nevertheless, this study has two major limitations. First, the number of CMD-affected patients was relatively small, although our sample size was relatively large among the published metagenomic studies on IBD. Large effects of gut dysbiosis were detected with strong statistical confidence in our study. However, it is likely that very small changes in microbial abundance and PRS in patients with CMD would not be detectable in our sample size. Second, the analyses of PRS and mbQTL were largely dependent on public resources generated from European populations. There is uncertainty regarding the cross-ancestry transferability of IBD PRS variants and the different genetic architectures of European and Korean populations in terms of allelic frequency and LD. Better analysis will be ensured by public resources generated from future East Asian-specific genome-wide association studies to identify mbQTLs and IBD susceptibility loci.

In summary, this study demonstrated how IBD-risk factors in the gut microbiota and human genetic variants are associated with prevalent comorbid depression and anxiety in patients with IBD through a series of comprehensive analyses in a large single-center prospective IBD cohort. The identified gut bacteria and genetic markers may serve as predictive biomarkers and therapeutic targets for mental disorders, thus providing new insights and opportunities to explore the underlying mechanisms of CMD development in IBD.

## Materials and methods

### Study cohort

All patients and healthy controls were enrolled at the IBD Center of Kyung Hee University Hospital (Seoul, Republic of Korea) between April 2018 and October 2020, after providing written informed consent. This study was approved by the Institutional Review Board of Kyung Hee University Hospital (KHUH 2018-03-006). The patients met the diagnostic criteria of CD or UC based on clinical, biochemical, stool, and endoscopic findings using histopathological and cross-sectional imaging methods.^42^ Clinical disease activity was measured using the Harvey-Bradshaw index (HBI) for CD and the partial Mayo score (PMS) for UC. Clinical remission was defined as HBI ≤ 4 and PMS ≤ 2.^43^

We obtained genomic and fecal microbial DNAs from genetically unrelated individuals over 17 years of age and collected basic demographic and clinical data from all participants (**Table 1**). Healthy controls had no serious illnesses, including gastrointestinal inflammation, and had no family history of IBD or mental disorders at enrollment. The fecal samples were collected using the Stool Nucleic Acid Collection and Preservation Tube (NORGEN BioTek) and promptly stored at −80 °C until DNA extraction.^44^

### Psychometric evaluation

The Hospital Anxiety and Depression Scale,^45^ a standardized psychological scale containing anxiety and depression subscales named HADS-A and HADS-D, respectively, was used for the psychometric testing of 225 patients with IBD. Significant anxiety and depression were defined as HADS-A ≥ 11 and HADS-D ≥ 11, respectively, as previously described.^46^

### Gut metagenome sequencing and microbial abundance analysis

The V3 to V4 regions of the 16S rRNA genes in fecal DNA were amplified using Agilent Herculase II fusion DNA Polymerase and Illumina Nextera XT library preparation kit v2 for paired-end sequencing on the MiSeq platform. The sequences of the targeted regions of the 16S rRNA genes were reconstructed by merging high-quality paired-end reads after trimming adapter bases and low-quality bases using Trimmomatic^47^ and QIIME2^48^. We employed an amplicon sequence variant (ASV) method^49^ to denoise sequence errors and remove chimeric sequences. Taxonomic classification and phylogenetic annotation were performed using reference sequences from Greengenes v13.8 database^50^.

We assessed the Shannon index, species richness by observed features, Simpson index, and Faith’s PD index to quantify the alpha diversity of the gut microbial community using library size-corrected metagenomic data. The beta diversity of the gut microbial composition among individual samples was examined using PCoA based on the Bray-Curtis dissimilarity of microbial abundance, which was normalized using DESeq2^51^. Differential beta diversity between sample groups of interest was evaluated by distance-based permutational multivariate analysis of variance using the adonis package^52^. Analysis of the differential abundance of the gut microbiota was performed using DESeq2 to estimate the log_2_-fold changes in abundance between sample groups. Sex, age, BMI, alcohol consumption, and current smoking status were adjusted in the case-control analysis. In the case-only differential abundance analysis, we additionally included IBD subtypes and disease activity scores as covariates.

### IBD MRS estimation

To understand the difference in the IBD-specific microbial burden between IBD patients with CMD and without CMD, we estimated the MRS for IBD based on the abundance of CMD-associated genera in each individual and the pre-estimated IBD-specific effect sizes of the same genera. The equation for the IBD MRS of an individual k (MRs_k_) is

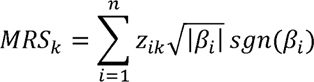

where *n* is the number of CMD-associated genera, *z_i_* is the normal transformed DESeq-normalized read count for the *i*-th genus in the individual *k*, and 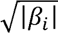 is the scaling factor to make *z_i_* have a variance as much as the absolute value of *β_i_* that is the pre-estimated log_2_-fold change in the abundance of the *i*-th genus in patients with IBD compared to healthy controls in the study. *sgn*(*β_i_*) is the sign function that returns +1 or −1 according to the sign of *β_i_*.

### Genotyping and whole genome imputation

Genome-wide data of single nucleotide polymorphisms (SNPs) in a subset of patients with IBD were generated using the Korea Biobank Array^53^ to estimate PRSs for IBD, depression, and anxiety in patients with or without CMD and to explore human mbQTLs for the bacteria of interest. After a general quality control procedure for the genotyping data (**Supplementary Table S1**), 541,449 high-quality variants with call rates ≥ 97%, minor allele frequencies ≥ 1%, and *P* values for Hardy-Weinberg equilibrium ≥ 1×10^−7^ were retained in 225 unrelated patients who showed a homogeneous genetic background and per-sample genotyping call rates ≥ 0.95. Whole genome imputation was performed using Minimac3^54^ with the reference panel derived from the 1000 Genomes Project phase 3 (1KGP3) data, after phasing the genotyping data into haplotypes using SHAPEIT software (v2.r904)^55^. All variants with imputation quality scores (R2) > 0.4 (n=10,689,279) were subjected to subsequent analyses.

### Genetic correlation

We estimated the between-trait genetic correlations of effect sizes of genome-wide variants on IBD, depression, and anxiety. We used the large-scale European GWAS summary statistics for IBD^27^, depression, and anxiety^28,29^, but not Asian GWAS statistics because our meta-analysis results for mbQTLs were mostly derived from European populations and there were no publicly available Asian GWAS summary statistics for the three traits. Genetic correlations were calculated using LDSC^56^ with pre-estimated scores of linkage disequilibrium (LD) in the 1KGP3 European population.

### PRS calculation

We calculated individual-level PRSs for IBD and mental disorders in IBD patients with HADS information based on the reported effect sizes of 136 variants for IBD (87 for UC, 119 for CD)^26,27^, 49 for depression, and 70 for anxiety^28,29^. The variants used for the PRS calculation were genetically and physically independent of each other (population-specific *r^2^*<0.2 and distance >1 Mb). They exceeded the GWAS significance threshold for disease risk in previous studies and are listed in our genetic data. Disease-specific PRS distributions in Korean individuals were obtained from the same variant sets in the whole genome imputation data of the Korean National Institute of Health cohort (n=72,179)^57^.

### MbQTL analysis for microbial taxa associated with both IBD and CMD

Genome-wide mbQTL analysis was performed to test for genetic associations with the abundance of the taxa of interest. The microbial abundance level of each taxon was normalized across samples using the rank-based inverse normal transformation. A multivariable linear regression model between microbial abundance and the allelic dosage of each genetic variant was examined using RVTESTS^58^, with adjustment for potential confounders such as sex, age, BMI, alcohol consumption, current smoking, IBD subtype, disease activity score, mental disorder status, and the top five genotypic principal components. A genome-wide mbQTL meta-analysis was performed using our result and a previous large-scale mbQTL analysis result^30^ (from 18,340 individuals; Europeans=78.0%, non-Europeans=22.0%) by METAL^59^ using the inverse variance-weighted fixed-effects model.

## Acknowledgements

This research was supported by a grant of the Korea Health Technology R&D Project through the Korea Health Industry Development Institute (KHIDI), funded by the Ministry of Health & Welfare, Republic of Korea (HI23C0661) and the Medical Research Center Program through the National Research Foundation (NRF) of Korea, funded by the Ministry of Science and ICT (NRF-2017R1A5A2014768).

## Data availability

All data relevant to the study are included in the article or uploaded as supplementary information. The other data are available from the corresponding authors upon reasonable request.

## Code availability

The analysis code from this paper can be obtained by contacting the corresponding author.

## Abbreviations used in this paper

ASV: amplicon sequence variant
BMI: body mass index
CD: Crohn’s disease
CI: confidence intervals
CMD: comorbid mental disorders
DAT: differentially abundant taxon
GWAS: genome-wide association study
HADS: hospital anxiety and depression scale
HBI: Harvey-Bradshaw index
IBD: inflammatory bowel disease
LD: linkage disequilibrium
mbQTL: microbial quantitative trait locus
MRS: microbial risk score
PC: principal component
PCoA: principal coordinate analysis
PD: phylogenetic diversity
PMS: partial Mayo score
PRS: polygenic risk score
SNP: single nucleotide polymorphism
UC: ulcerative colitis
1KGP3: 1000 Genomes Project phase 3

