## Supplementary Figure 1-4 for "Gut microbial and human genetic signatures of inflammatory bowel disease increase risk of comorbid mental disorders"

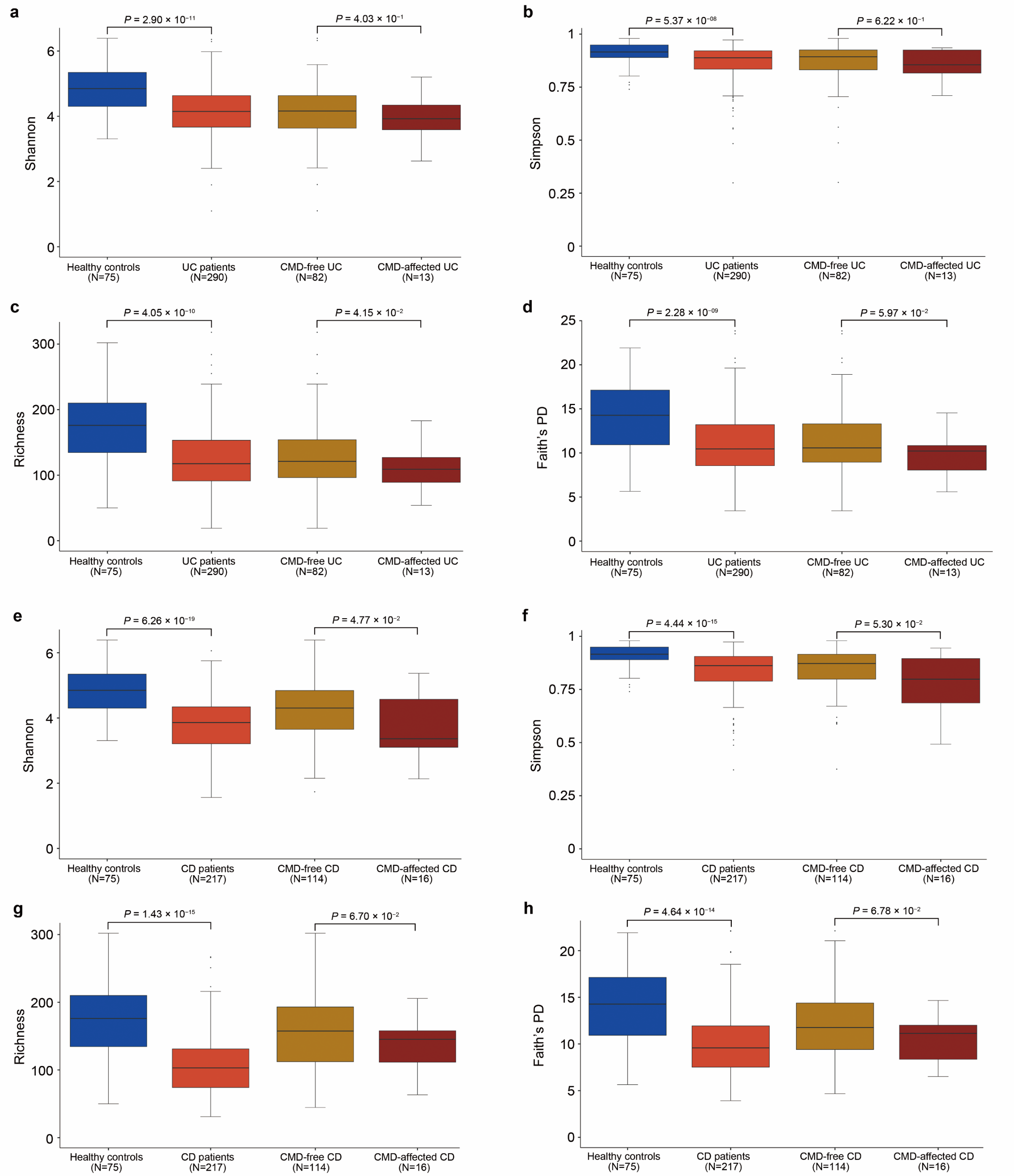

**Supplementary Figure S1. Alpha diversity distribution stratified by UC and CD.**

**a-h,** the patients with IBD were divided into UC (**a**-**d**) and CD (**e***-***h**). Alpha diversity was assessed based on Shannon index (**a**, **e**), Simpson index (**b**, **f**), richness (**c**, **g**), and Faith’s PD index (**d**, **h**) at the genus level.

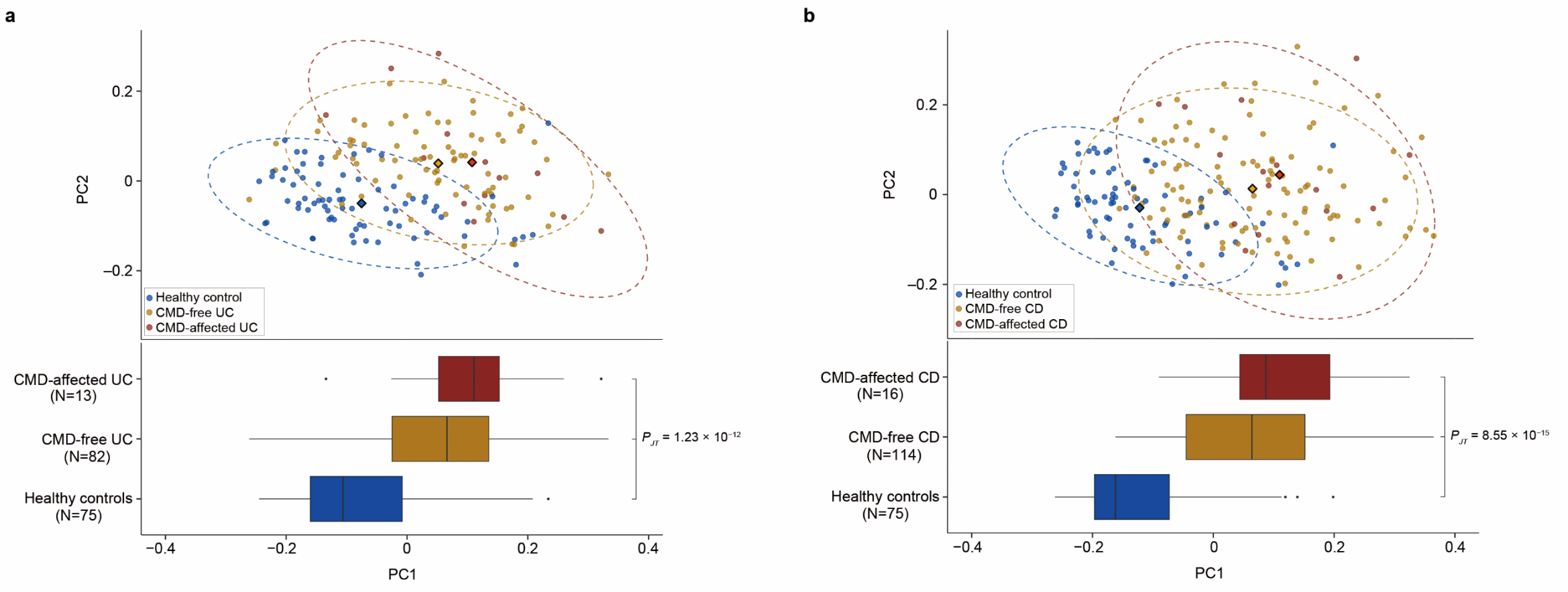

**Supplementary Figure S2. Beta diversity distribution stratified by UC and CD.**

Beta diversity was measured based on the Bray-Curtis dissimilarity of individuals’ genus abundance and visualized in a PCoA plot according to the top two principal components (PCs) for UC (**a**) and CD (**b**) separately. The centroid of each cluster is marked by a diamond. The top PC that explained the largest proportion of variance in microbial composition in the study subjects was shown in a boxplot according to the affected status. The p-value from a Jonckheere-Terpstra test (*P*_JT_) was calculated to test for a significant trend toward higher PC1 values in the order of healthy controls, CMD-free patients, and CMD-affected patients.

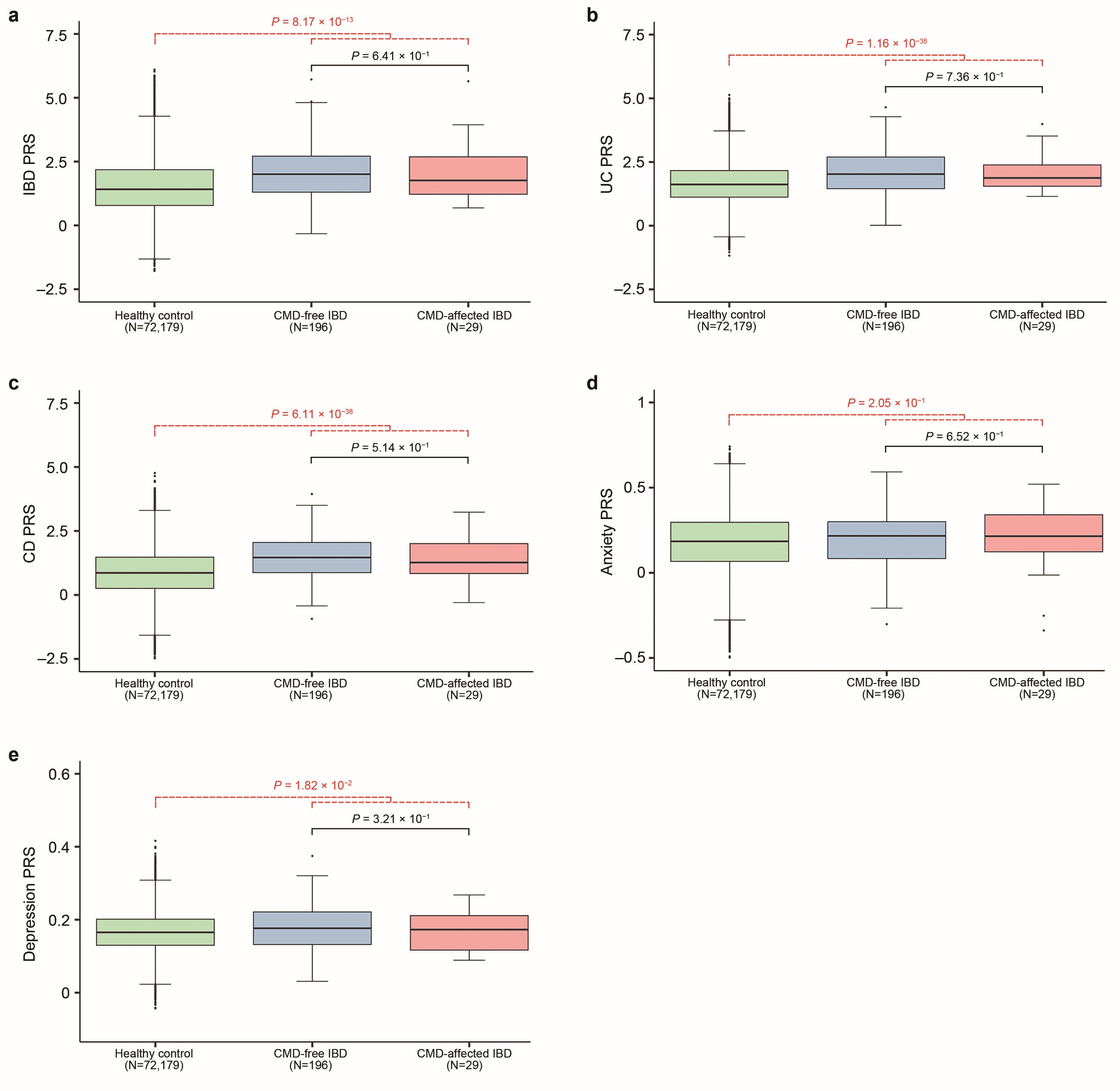

**Supplementary Figure S3. Distributions of PRSs for IBD, UC, CD, anxiety, and depression.**

**a-e,** PRSs for IBD (**a**), UC (**b**), CD (**c**), anxiety (**d**), and depression (**e**) were calculated in the out-of-study healthy controls, CMD-free IBD patients and CMD-affected IBD patients. *P* values for the difference in PRSs between healthy controls and IBD patients as well as between CMD-free and CMD-affected IBD patients were calculated by Wilcoxon rank sum tests and are represented in red and black colors, respectively.

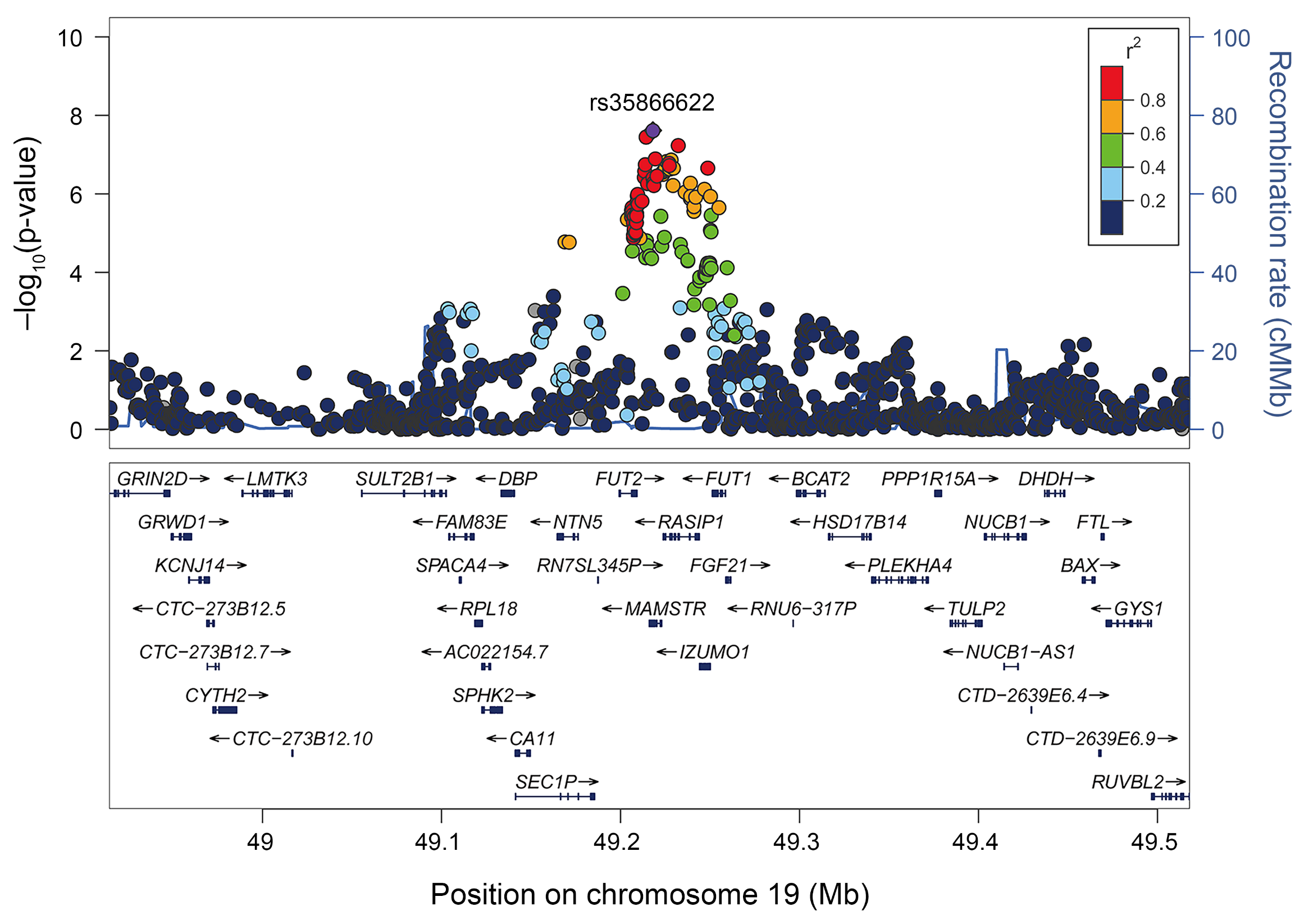

**Supplementary Figure S4. An IBD-associated mbQTL for the genus *Ruminococcus* (*torques* group).**

A *P*_meta_ value for the association between the abundance of the genus *Ruminococcus* (*torques* group) and each variant was plotted in a minus log_10_ scale according to the physical position (hg19) of the variants. The *T* allele of the lead mbQTL variant rs35866622, a known IBD-associated variant, was associated with a decrease in the abundance of the genus *Ruminococcus* (*torques* group) (*P*_meta_=2.43x10^−8^; β=−0.061; standard error=0.011).
