## Supplementary Note for "Gut microbial and human genetic signatures of inflammatory bowel disease increase risk of comorbid mental disorders"

**Supplementary Note 1. Short review on IBD/CMD-associated genera**

We identified that nine genera were associated with both IBD and CMD, exhibiting the same directional changes in abundance. The pathological and psychological effects of these genera on human diseases including inflammatory and mental disorders have poorly understood. After a literature review, we briefly summarize the most relevant reports on those genera of interest (excluding three unclassified genera and two genera that we already described in the main text), especially regarding their metabolites and the associations with diseases.

1. *Leptotrichia*:

Some species of the genus *Leptotrichia* living in several tracts of human, mostly in mouth, have been considered as a trigger of dysbiosis to mediate IBD.^1^ The increased abundance of *Leptotrichia* was detected in IBD patients,^1^ showing the same direction seen in our case-control differential abundance analysis. This genus is known for generating H_2_S through the fermentation of amino acids containing sulfur. The H_2_S-high producing bacteria have been correlated with the severity of inflammation in CD patients.^2^ In addition, *Leptotrichia* sp. was positively associated with the severity of depression in patients with schizophrenia.^3^

2. *Moryella*:

The genus *Moryella* produces anti-inflammatory metabolites including acetate, butyrate and indole that are also known to promote gut homeostasis.^4, 5^ A gut metagenomic analysis in patients with Parkinson’s disease suggested the involvement of *Moryella* in the gut-brain axis.^6^ The abundance of the genus was decreased after treatment for Parkinson’s disease, indicating that the increased abundance *Moryella* could be associated with risk of the neurological disorder. Similarly, we observed the increased abundance in CMD-affected IBD patients.

3. *Shuttleworthia*:

*Shuttleworthia* is a gram-positive and non-motile genus with a single known species from human oral cavity that produces acetate, butyrate and lactate as metabolites.^7^ We observed lowered abundances in CMD-affected IBD patients. In contrast, an opposite direction of the abundance change has been reported in individuals with anxiety and depression symptoms in a general population,^8^ although other recent studies showed no evidence of differential abundance changes in *Shuttleworthia*.^9-12^

4. *Haemophilus*:

The genus *Haemophilus* includes many species to mediate a variety of infectious diseases (e.g., *Haemophilus* *influenzae*).^13^ This genus resides on the mouth, vagina, upper respiratory tract, and gastrointestinal tract.^14^ In a previous study,^15^ the genus *Haemophilus* was significantly lower in IBD patients compared to healthy controls in a salivary microbial analysis. In addition, several studies reported the decreased level of *Haemophilus* in individuals with psychiatric disorders.^16^ All these observations were consistent with lowered abundances in patients with IBD and CMD in our study.
